# The effects of oxygenation on acute vasodilator challenge in pulmonary arterial hypertension

**DOI:** 10.1101/2023.04.27.23289235

**Authors:** Matthew D Rockstrom, Ying Jin, Ryan A Peterson, Peter Hountras, David Badesch, Sue Gu, Bryan Park, John Messenger, Lindsay M. Forbes, William K. Cornwell, Todd M Bull

## Abstract

**Background:** Identification of long-term calcium channel blocker (CCB) responders with acute vasodilator challenge is critical in the evaluation of patients with pulmonary arterial hypertension. Currently there is no standardized approach for use of supplemental oxygen during acute vasodilator challenge.

**Methods:** Retrospective analysis of patients identified as acute vasoresponders, treated with CCBs. All patients had hemodynamic measurements in three phases: 1) at baseline; 2) with 100% fractional inspired oxygen; and 3) with 100% fractional inspired oxygen plus inhaled nitric oxide (iNO). Patients were divided into two cohorts. Those meeting the definition of acute vasoresponsiveness from phase 2 to phase 3 were labeled “iNO Responders.” Those who did not reach the threshold of acute vasoresponsiveness from phase 2 to phase 3 but did meet the definition from phase 1 to phase 3 were labeled “Oxygen Responders.” Survival, hospitalization for decompensated right heart failure, duration of CCB monotherapy, and functional data were collected.

**Results:** iNO Responders, when compared to Oxygen Responders, had superior survival (100% vs 50.1% 5-year survival, respectively), fewer hospitalizations for acute decompensated right heart failure (0% vs 30.4% at 1 year, respectively), longer duration of CCB monotherapy (80% versus 52% at 1 year, respectively), and superior six-minute walk distance.

**Conclusion:** Current guidelines for acute vasodilator testing do not standardize oxygen coadministration with iNO. This study demonstrates that adjusting for the effects of supplemental oxygen before assessing for acute vasoresponsiveness identifies a cohort with superior functional status, tolerance of CCB monotherapy, and survival while on long-term CCB therapy.

---

Pulmonary arterial hypertension (PAH) is a progressive disease of the small pulmonary arteries characterized by increased pulmonary vascular resistance and eventual right heart failure (1). Despite advancements in treatment, PAH is a life-limiting and often terminal disease (2, 3). A subset of 5 – 10% of PAH patients demonstrate excellent response to long-term calcium channel blockers (CCB) therapy (4). Recognition of long-term CCB responders is essential for selection of appropriate therapy and prognostication as these patients have less severe functional impairments, improved hemodynamics, and significantly prolonged life span on CCB monotherapy (5, 6). However, in unresponsive patients, CCB use is contraindicated and can cause hemodynamic decompensation and increased mortality (6), making correct identification of this population imperative.

Long-term CCB responders are identified by acute vasodilator challenge during right heart catheterization (RHC) as part of the initial workup of PAH (7). Identification of acute vasoresponders requires a reduction in mean pulmonary arterial pressure (mPAP) of ≥ 10 mmHg reaching an absolute mPAP ≤ 40 mmHg without a decrease in cardiac output (CO) in response to an acute vasodilatory agent (6, 8). Inhaled nitric oxide (iNO) is an endogenous vasodilator produced by vascular endothelial cells and is frequently used to assess acute vasoreactivity in PAH (9, 10). iNO can be delivered at variable concentrations and is often co-administered with supplemental oxygen, however there is significant variability in the use of supplemental oxygen during acute vasodilator challenge, ranging from maintenance of fractional inspired oxygen (FiO2) of 21% to use of 100% FiO2 administered with iNO (11-15). The effects of supplemental oxygen are not inert, as alveolar hypoxia has long been understood to induce pulmonary vasoconstriction and subsequent elevation in mPAP (16-18) and increased alveolar oxygen tension reduces mPAP (19, 20) and pulmonary vascular resistance (PVR) (21) in PAH patients.

In this study, we evaluated the long-term tolerance of CCBs amongst two cohorts – “iNO Responders,” who meet the definition of acute vasoresponsiveness only after first adjusting for the effects of supplemental oxygen, and “Oxygen Responders,” who meet the definition of acute vasoresponsiveness without adjusting for the vasodilatory effects of oxygen. For each cohort, we follow tolerance of CCB monotherapy, mortality, hospitalization for decompensated right heart failure, and functional status following the initiation of CCB.

## Materials and Methods

### Study group

We retrospectively evaluated medical records of all patients, aged 18 or older, referred to a large regional tertiary pulmonary hypertension care center (PHCC) for management of their PAH from January 2010 to October 2021. The study was approved by the Colorado Multiple Institutional Review Board (COMIRB Protocol #: 07-0953).

PAH was defined by an mPAP ≥ 25 mmHg with a pulmonary capillary wedge pressure (PCWP) of < 15 mmHg and pulmonary vascular resistance (PVR) > 3 Woods units in the absence of chronic thromboembolic pulmonary hypertension or significant chronic respiratory disease (22). All PAH patients were evaluated and patients were identified as vasoresponders if they 1) met the definition of acute vasoresponsiveness, with a reduction in mPAP ≥ 10 mmHg and a final mPAP ≤ 40 mmHg (23); 2) were initially treated with CCB monotherapy for their PAH; and 3) acute vasodilator challenge included a dedicated measurement of mPAP during an “oxygen phase” at 100% FiO_2_, in addition to hemodynamic measurements at baseline and “vasodilator phase” with 100% FiO_2_ and 40ppm iNO. In accordance with the American College of Chest Physicians recommendations (24) for evaluation of acute vasoreactivity in pulmonary hypertension, all World Health Organization (WHO) Group I patients (25) were eligible for enrollment including: idiopathic PAH (iPAH), familial PAH (FPAH) and PAH associated with connective tissue disease, portopulmonary hypertension, toxins, congenital heart disease, and HIV. We also performed a subgroup analysis of patients diagnosed with iPAH, FPAH, and toxin-mediated PAH, in line with the 6^th^ World Symposium on Pulmonary Hypertension (WSPH) and European Society of Cardiology and European Respiratory Sociate (ECS/ERS) recommendations (26, 27) for acute vasoreactivity challenge. WHO class was determined by a clinician expert in the treatment of pulmonary vascular disease and re-adjudicated by two authors (MR, TB).

### Right heart catheterization and acute vasodilator challenge

All right heart catheterizations were performed using standard technique as previously described (13). Baseline hemodynamic measurements including right atrial pressure (RAP), mPAP, PWCP, and CO were first recorded on ambient room air. If the patient had chronic oxygen requirements, baseline measurements were recorded at their home supplemental oxygen dose with a goal saturation of peripheral oxygen (SpO_2_) of 90-94%. mPAP was recorded during an intermediate “oxygen phase,” using 100% supplemental oxygen via non-rebreather face mask for 2 minutes. Finally, complete hemodynamic measurements were repeated during a final “vasodilator phase,” following administration of 40 ppm of iNO and 100% supplemental oxygen for 5 minutes.

### Responder definitions, CCB administration, and monitoring

Patients were designated as “iNO Responders” if they met the definition of acute vasoresponsiveness from the effects of iNO after first normalizing for the effects of supplemental oxygen. If patients met the definition of acute vasoresponsiveness only through a combination of the effects of oxygen and iNO, they were labeled “Oxygen Responders” (*Figure 1*).

**Fig 1:**
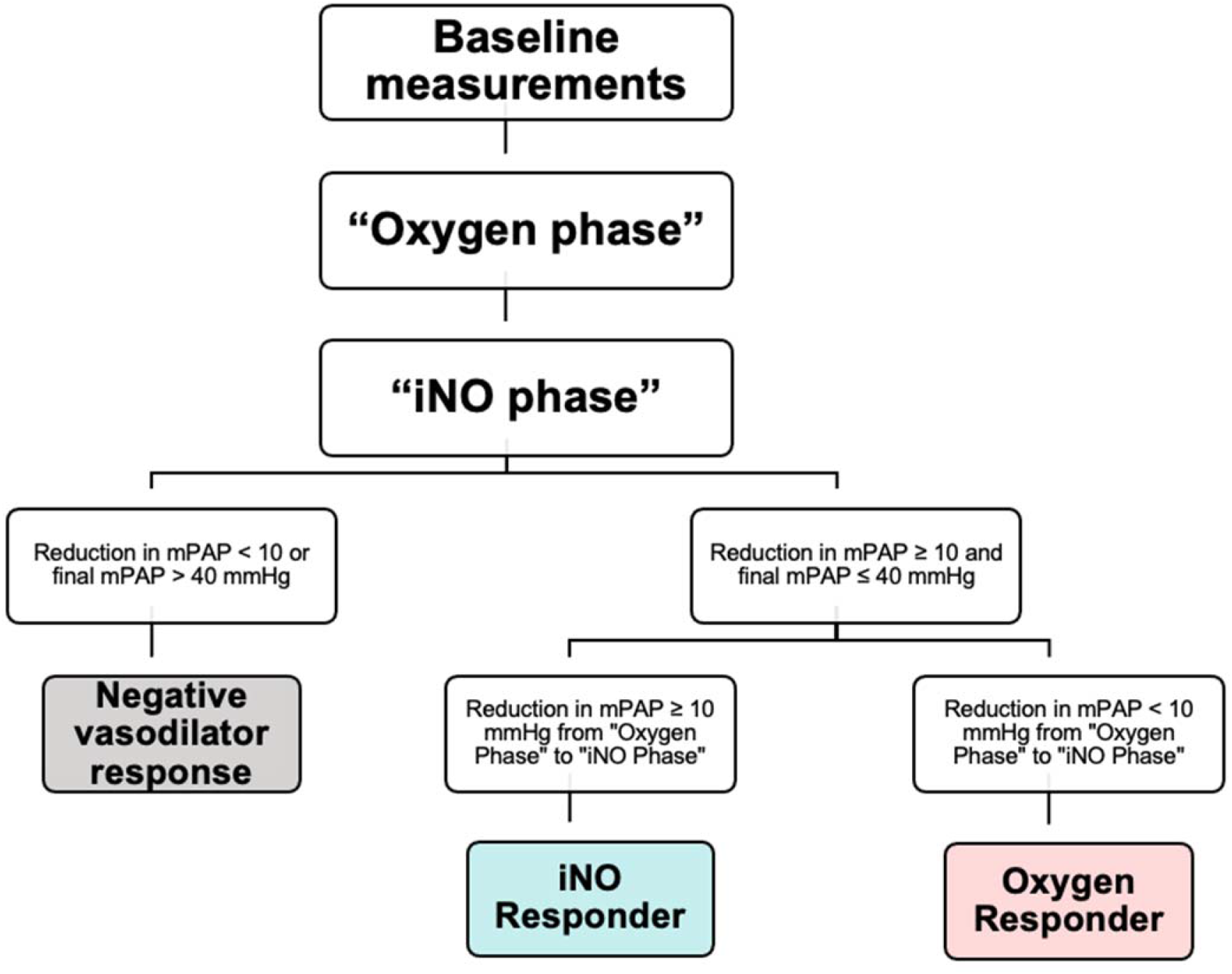
Definitions of iNO and Oxygen Responders. Mean pulmonary arterial pressure (mPAP) was measured at baseline (ambient room air or patient’s chronic home oxygen requirement, if applicable), after 2 min at 100% fractional inspired oxygen (“Oxygen Phase”), and after 5 min at 100% FiO_2_ + 40 ppm inhaled nitric oxide (“Vasodilator Phase”). Both iNO and Oxygen responders had a reduction of 10 mmHg from baseline to Vasodilator Phase with a final mPAP ≤ 40 mmHg. “iNO Responders” had a 10 mmHg reduction in mPAP from Oxygen Phase to Vasodilator Phase. “Oxygen Responders” had a < 10 mmHg reduction in mPAP from Oxygen Phase to Vasodilator Phase.

Selection and titration of a CCB agent was performed at the discretion of the treating pulmonary hypertension provider. CCB monotherapy was discontinued if the patient did not have favorable functional response: improvement to NYHA FC I or II, a lack of improvement in 6MWD, or if they had evidence of worsening mPAP or right ventricular function on transthoracic echocardiogram or RHC. NYHA functional status and 6MWD were collected by chart review at baseline, 1-, and 3-year follow-up. Hospitalizations for acute decompensated right heart failure within the first year of initiation of CCB were observed. Additionally, time on CCB monotherapy and mortality from time of initiation of CCB were collected.

### Statistical analysis

For demographic variables, we presented summary statistics stratified by vasoresponsive status alongside non-parametric statistical tests (Kruskal-Wallis tests for continuous variables, Fisher’s exact test for categorical variables). For time-to-event outcomes, we presented stratified Kaplan-Meier estimates of event-free probabilities, as well as *P* values from log-rank tests. We computed restricted means for each event time for additional descriptive statistics as the median event time was not observed for all outcomes. For the longitudinal outcome of 6MWD, we presented summary statistics and visualizations of means and standard deviations stratified by vasoresponsive status. Differences at each time were assessed using a linear mixed model with subject-level random intercepts and fixed effects for time, vasoresponsive status, and their interaction. Using the mixed model for 6MWD, group-level differences at each time were tested using t-tests with the Kenward-Roger approximation for degrees of freedom, while differences in NYHA classification were tested separately at each time point using Fisher’s Exact tests.

## Results

Two hundred sixty eight consecutive PAH patients were identified. Of these, 91 patients (30.3%) had iPAH, 3 (1.0%) had familial PAH, and 206 (68.7%) had associated PAH. A total of 33 acute vasoresponders were identified. Amongst these, 10 were “iNO Responders” and 23 were “Oxygen Responders”. Average duration of follow up was 4.5 years. Basic demographic characteristics and rates of comorbid systemic hypertension, type 2 diabetes mellitus, and smoking history were similar between the two cohorts. Eight iNO Responders and 16 Oxygen Responders had chronic oxygen requirements (80% versus 69.6%, respectively, *P* = 0.037). Amongst iNO Responders, 7 (70.0%) had iPAH, 2 (20.0%) had CTD-associated PAH, and 1 (10.0%) had FPAH. Amongst Oxygen Responders, 6 (26.1%) had iPAH and 17 (73.9%) had associated PAH, including 8 (34.8%) with CTD, 4 (17.4%) toxin associated, 3 (13.0%) with congenital heart disease. 2 (8.7%) with portopulmonary disease (*Table 1*).

**Table 1:**
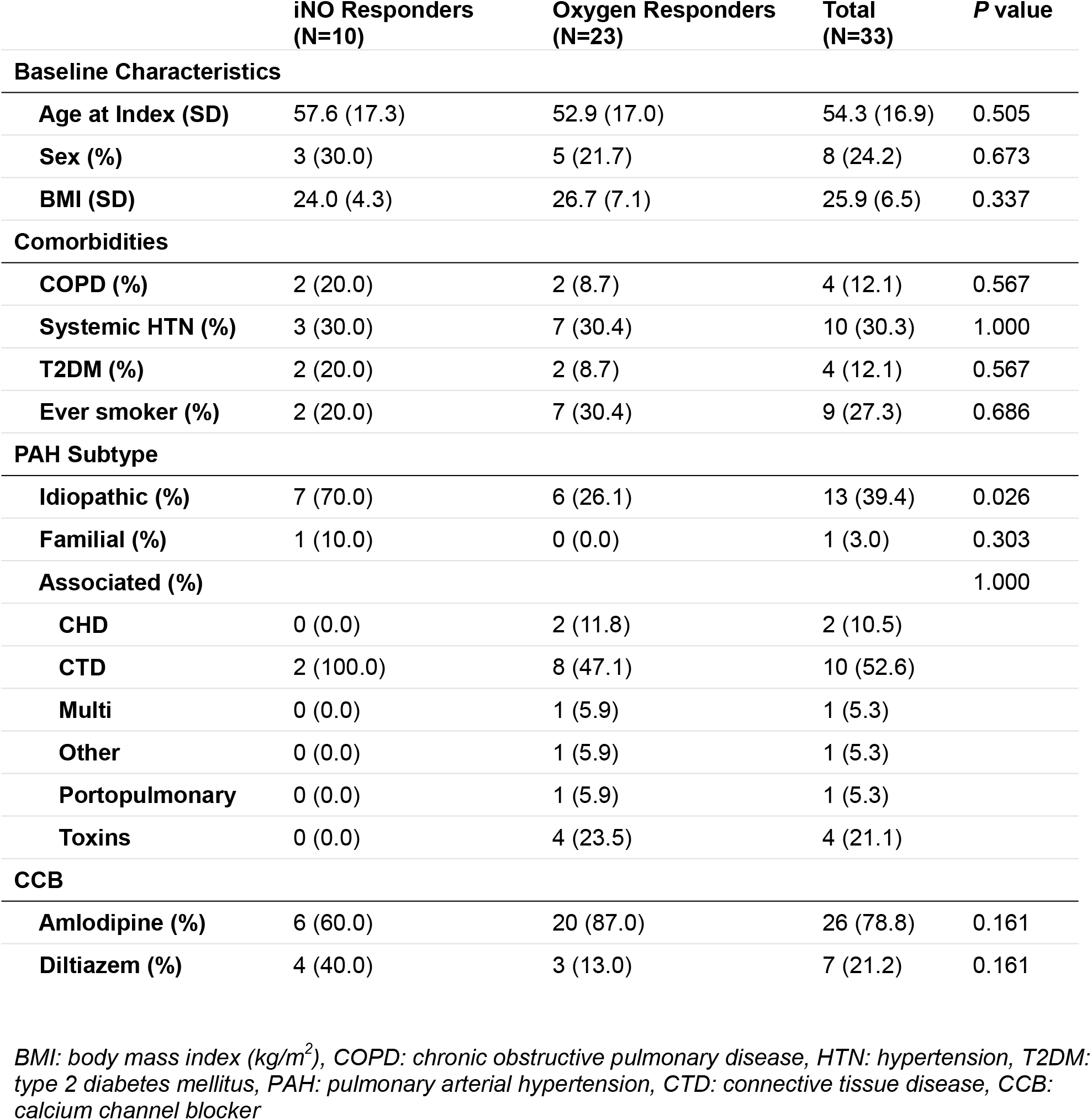
Baseline Clinical Characteristics and Calcium Channel Blocker Use of Study Cohort.

|  | iNO Responders<br>(N=10) | Oxygen Responders<br>(N=23) | Total<br>(N=33) | P value |
| --- | --- | --- | --- | --- |
| **Baseline Characteristics** |  |  |  |  |
| **Age at Index (SD)** | 57.6 (17.3) | 52.9 (17.0) | 54.3 (16.9) | 0.505 |
| **Sex (%)** | 3 (30.0) | 5 (21.7) | 8 (24.2) | 0.673 |
| **BMI (SD)** | 24.0 (4.3) | 26.7 (7.1) | 25.9 (6.5) | 0.337 |
| **Comorbidities** |  |  |  |  |
| **COPD (%)** | 2 (20.0) | 2 (8.7) | 4 (12.1) | 0.567 |
| **Systemic HTN (%)** | 3 (30.0) | 7 (30.4) | 10 (30.3) | 1.000 |
| **T2DM (%)** | 2 (20.0) | 2 (8.7) | 4 (12.1) | 0.567 |
| **Ever smoker (%)** | 2 (20.0) | 7 (30.4) | 9 (27.3) | 0.686 |
| **PAH Subtype** |  |  |  |  |
| **Idiopathic (%)** | 7 (70.0) | 6 (26.1) | 13 (39.4) | 0.026 |
| **Familial (%)** | 1 (10.0) | 0 (0.0) | 1 (3.0) | 0.303 |
| **Associated (%)** |  |  |  | 1.000 |
| **CHD** | 0 (0.0) | 2 (11.8) | 2 (10.5) |  |
| **CTD** | 2 (100.0) | 8 (47.1) | 10 (52.6) |  |
| **Multi** | 0 (0.0) | 1 (5.9) | 1 (5.3) |  |
| **Other** | 0 (0.0) | 1 (5.9) | 1 (5.3) |  |
| **Portopulmonary** | 0 (0.0) | 1 (5.9) | 1 (5.3) |  |
| **Toxins** | 0 (0.0) | 4 (23.5) | 4 (21.1) |  |
| **CCB** |  |  |  |  |
| **Amlodipine (%)** | 6 (60.0) | 20 (87.0) | 26 (78.8) | 0.161 |
| **Diltiazem (%)** | 4 (40.0) | 3 (13.0) | 7 (21.2) | 0.161 |

*BMI: body mass index (kg/m<sup>2</sup>), COPD: chronic obstructive pulmonary disease, HTN: hypertension, T2DM: type 2 diabetes mellitus, PAH: pulmonary arterial hypertension, CTD: connective tissue disease, CCB: calcium channel blocker*

### Hemodynamics

Baseline hemodynamic measurements between iNO Responders and Oxygen Responders revealed similar baseline RAP, mPAP, PCWP, and PVR. Amongst iNO Responders, there was lower mean baseline CO than Oxygen Responders (4.0 +/- 1.1 versus 5.6 +/- 2.1 L/min, respectively, *P* = 0.010). Oxygen Responders trended towards lower mPAP than iNO Responders during oxygen phase (35.8 versus 44.3 mmHg, respectively, *P* = 0.057) and both iNO Responders and Oxygen Responders had similar mPAP during vasodilator phase (30.0 +/- 7.7 versus 29.8 +/- 7.4 mmHg, *P* = 0.89). (*Table 2*). Oxygen Responders had a greater reduction in mPAP from baseline to oxygen phase than iNO Responders (8.0 versus 4.3 mmHg, respectively, *P* = 0.021) and iNO Responders’ reduction in mPAP was similar to that seen in the overall cohort of 268 PAH patients (3.9 mmHg). iNO Responders had a greater reduction in mPAP from oxygen phase to vasodilator phase than Oxygen Responders (14.4 versus 6.0 mmHg, respectively, *P* < 0.001) (*Figure 2*).

**Table 2:** Hemodynamic Data for Study Cohort during Right Heart Catheterization.

|  | iNO Responders (N=10) | Oxygen Responders (N=23) | P value |
| --- | --- | --- | --- |
| **Baseline** |  |  |  |
| **RAP, mmHg (SD)** | 5.10 (3.54) | 5.78 (3.00) | 0.568 |
| **mPAP, mmHg (SD)** | 48.70 (13.62) | 43.83 (7.82) | 0.398 |
| **PCWP, mmHg (SD)** | 7.90 (3.67) | 9.52 (3.70) | 0.243 |
| **CO,TD L/min (SD)** | 3.97 (1.11) | 5.61 (2.15) | 0.010 |
| **CO,Fick L/min (SD)** | 4.04 (1.06) | 4.91 (1.15) | 0.051 |
| **PVR, WU (SD)** | 11.03 (5.85) | 8.14 (3.47) | 0.177 |
| **Oxygen Phase** |  |  |  |
| **mPAP, mmHg (SD)** | 44.40 (11.72) | 35.83 (7.72) | 0.057 |
| **Vasodilator Phase** |  |  |  |
| **RAP, mmHg (SD)** | 4.60 (2.91) | 4.83 (3.20) | 0.735 |
| **mPAP, mmHg (SD)** | 30.00 (7.73) | 29.83 (7.43) | 0.890 |
| **PCWP, mmHg (SD)** | 8.00 (3.86) | 10.61 (4.12) | 0.051 |
| **CO,TD L/min (SD)** | 4.17 (1.22) | 5.10 (1.77) | 0.183 |
| **PVR, WU (SD)** | 5.55 (2.49) | 3.89 (2.07) | 0.044 |

All figures listed with standard deviations in parentheses; RAP: right atrial pressure; mPAP: mean pulmonary arterial pressure; PCWP: pulmonary capillary wedge pressure; CO: cardiac output; TD: thermodilution; PVR: pulmonary vascular resistance; WU: Woods Units "Oxygen Phase" with hemodynamic measurements taken after 100% supplemental oxygen for 2 minutes; "Vasodilator Phase" with hemodynamic measurements take after 100% supplemental oxygen + 40 ppm iNO for 5 min

**Fig 2.**
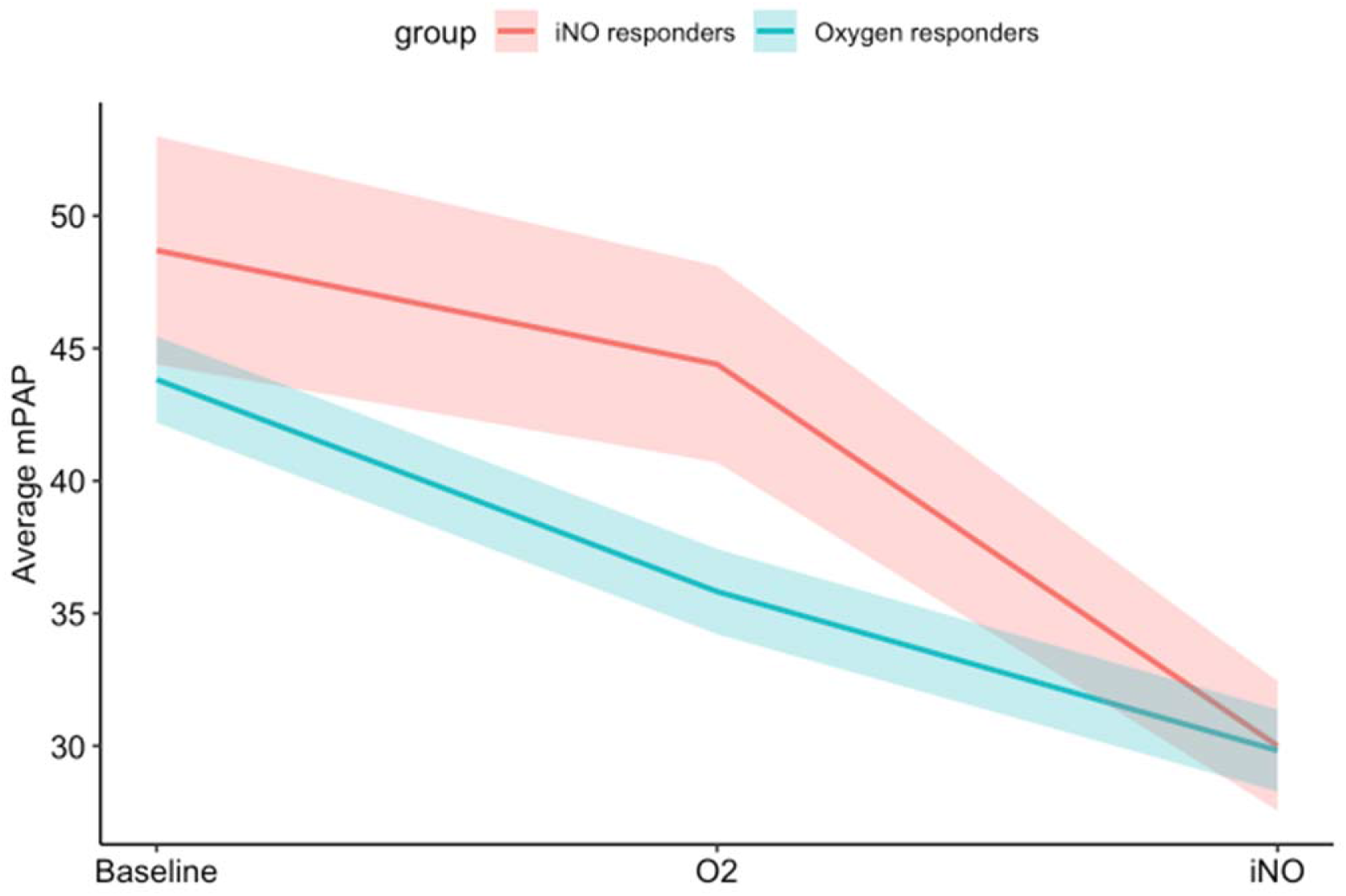
Mean pulmonary arterial pressure (mPAP) for iNO Responders (shown in red) and Oxygen Responders (shown in blue) measured at baseline, following administration of 100% FiO_2_ (indicated as “O2”), and following administration of 100% FiO_2_ with 40 ppm iNO (indicated as “iNO”). Ribbons represent the average mPAP +/- 1 standard error, calculated separately from data at each time point

### Survival, hospital free survival, CCB monotherapy

iNO Responders had superior survival and hospital-free survival relative to Oxygen Responders (*Figure 3*). There were no deaths within 10 years of initiation of CCB amongst iNO Responders. Amongst Oxygen Responders, survival was 87.0% at one year, 71.6% at 3 years, and 50.1% at 5 years from initiation of CCB. Additionally, there were no hospitalizations for acute decompensated right heart failure within a year of initiation of CCB amongst iNO Responders as compared to 7 (30.4%) amongst Oxygen Responders. iNO Responders had greater tolerance of CCB monotherapy compared to Oxygen Responders (*Figure 4*). At one year follow up, 80% of iNO Responders were still on monotherapy as compared to 52% of Oxygen Responders. Restricted 10-year mean time of CCB monotherapy was 6.0 years versus 2.8 years in iNO Responders and Oxygen Responders, respectively (log rank *P* = 0.0085).

**Fig 3.**
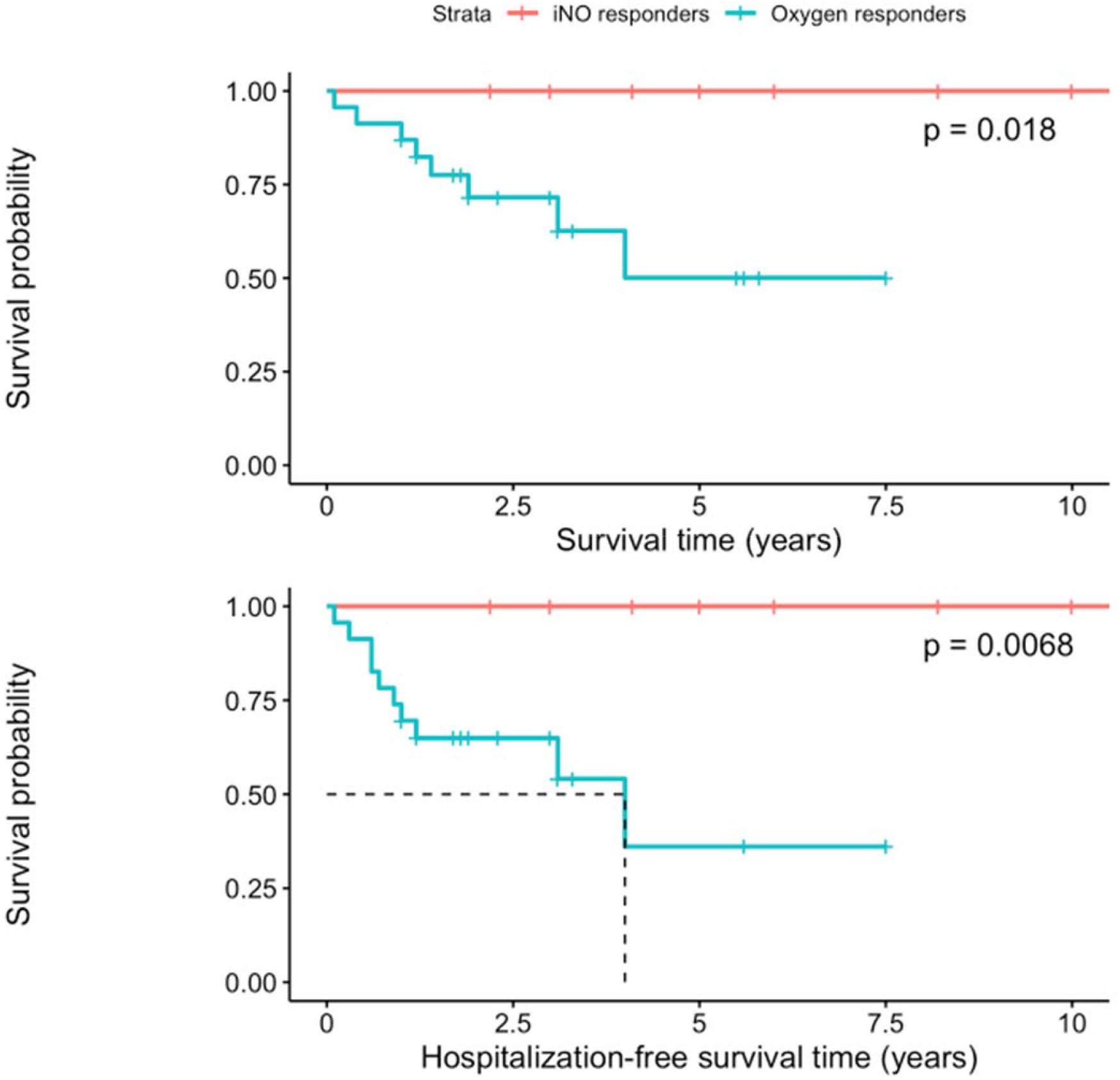
Kaplan-Meier curves for survival (above) and hospitalization-free survival (below) following initiation of calcium channel blocker therapy for 33 patients, subdivided into iNO Responders (n = 10 at initiation, shown in red) and Oxygen Responders (n = 23 at initiation, shown in blue). Dotted lines indicate mean survival time and hospitalization-free survival time, respectively. Restricted mean 10-year survival time amongst Oxygen Responders was 8.59 years and was 6.71 years for hospitalization-free survival. All relationships were statistically significant.

**Fig 4.**
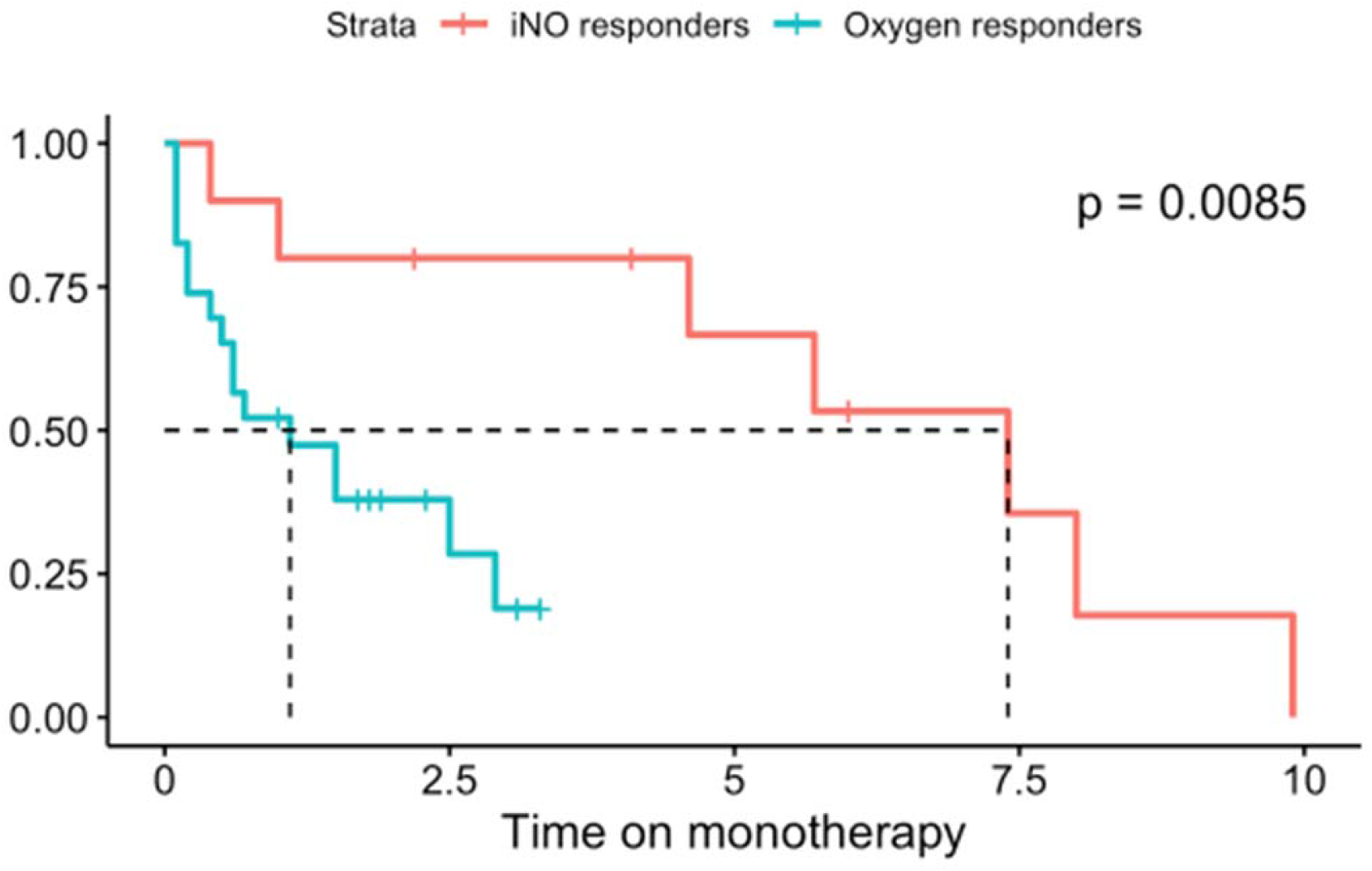
Kaplan-Meier curve for calcium channel blocker monotherapy amongst 33 patients, subdivided into iNO Responders (n = 10 at initiation) and Oxygen Responders (n = 23 at initiation). Dotted lines indicate mean duration of monotherapy for both cohorts. Restricted mean 10-year duration of monotherapy for iNO Responders was 6.01 years and for Oxygen Responders was 2.76 years.

A subgroup analysis of patients with iPAH, FPAH, and toxin-mediated PAH demonstrated that iNO Responders had longer survival time (*P* = 0.009) and hospital-free survival time (*P =* 0.013) than Oxygen Responders (*Figure 5*). Additionally, iNO Responders had longer time on monotherapy than Oxygen responders (7.4 versus 1.5 years, respectively, *P* = 0.085).

**Fig 5.**
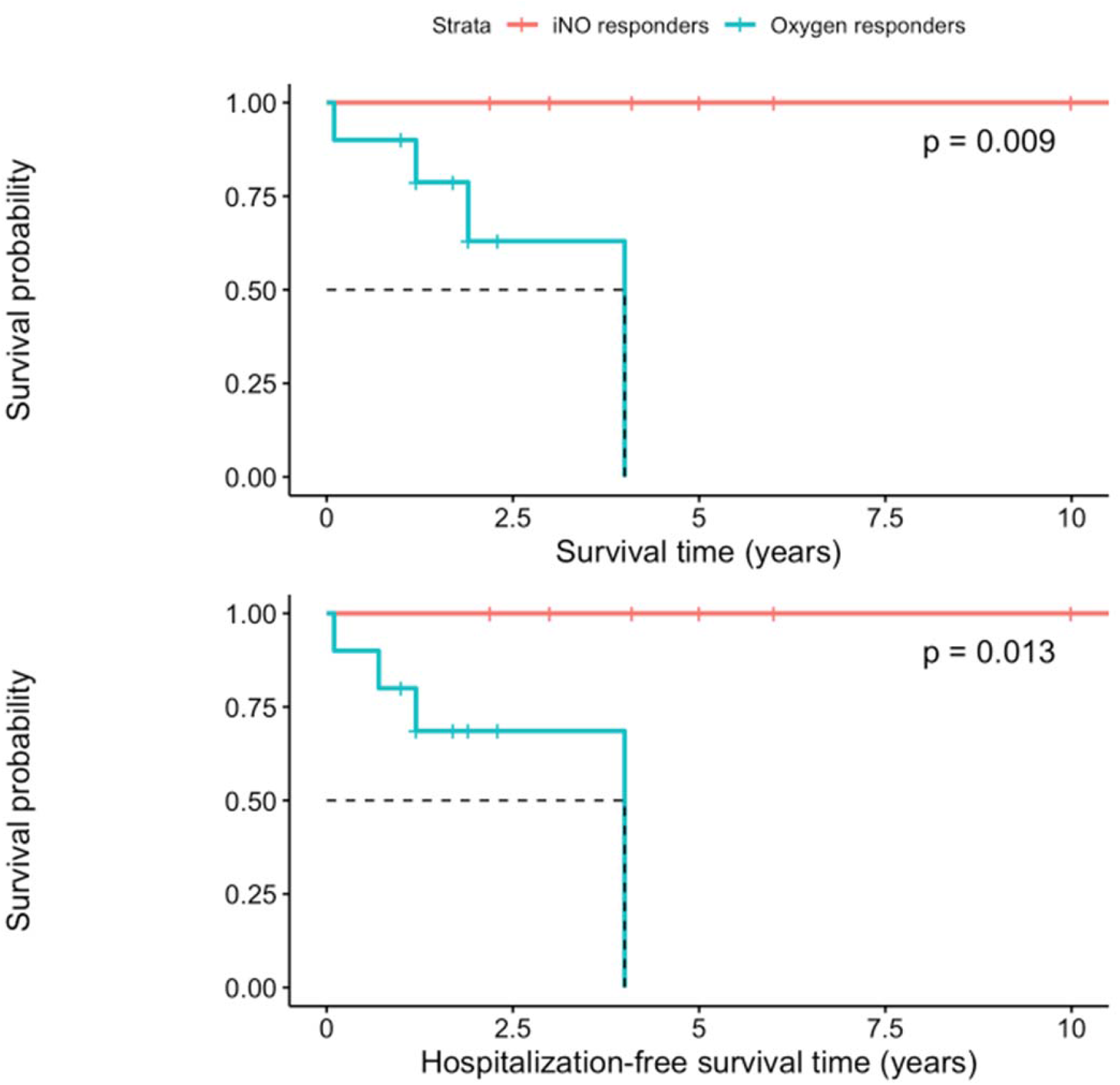
Kaplan-Meier curves for survival (above) and hospitalization-free survival (below) following initiation of calcium channel blocker therapy for subgroup analysis of patients meeting recommendations for acute vasoresponsiveness screening per 6^th^ World Symposium of Pulmonary Hypertension (WSPH) and European Society of Cardiology/European Respiratory (ERC/ERS) guidelines. Dotted lines indicate mean survival time and hospitalization-free survival time, respectively.

### Functional status

The 6MWD of iNO Responders and Oxygen Responders were 423 versus 352 meters at baseline (*P* = 0.45), 409 versus 292 meters at 1-year (*P* = 0.21), and 454 versus 224 meters at 3-years, respectively (*P* = 0.017) (*Figure 6*). iNO Responders trended towards superior functional class at 1- and 3-year follow up, although this did not reach the threshold for statistical significance (*Table 3*).

**Table 3:** New York Heart Association Functional Class for Studied Cohort at Baseline, 1-year, and 3-year Follow Up.

|  | iNO Responders (N=10) | Oxygen Responders (N=23) | P value |
| --- | --- | --- | --- |
| **Baseline** |  |  | 0.678 |
| I | 1 (11.1%) | 3 (16.7%) |  |
| II | 7 (77.8%) | 9 (50.0%) |  |
| III | 1 (11.1%) | 5 (27.8%) |  |
| IV | 0 (0.0%) | 1 (5.6%) |  |
| **Year 1** |  |  | 1.000 |
| I | 2 (40.0%) | 1 (20.0%) |  |
| II | 2 (40.0%) | 2 (40.0%) |  |
| III | 1 (20.0%) | 2 (40.0%) |  |
| **Year 3** |  |  | 0.679 |
| I | 2 (40.0%) | 0 (0.0%) |  |
| II | 2 (40.0%) | 1 (33.3%) |  |
| III | 1 (20.0%) | 2 (66.7%) |  |

*New York Heart Association classes I – IV (indicated as I, II, III, and IV) recorded at time of right heart catheterization and again at one-year and three-year follow up*

**Fig 6.**
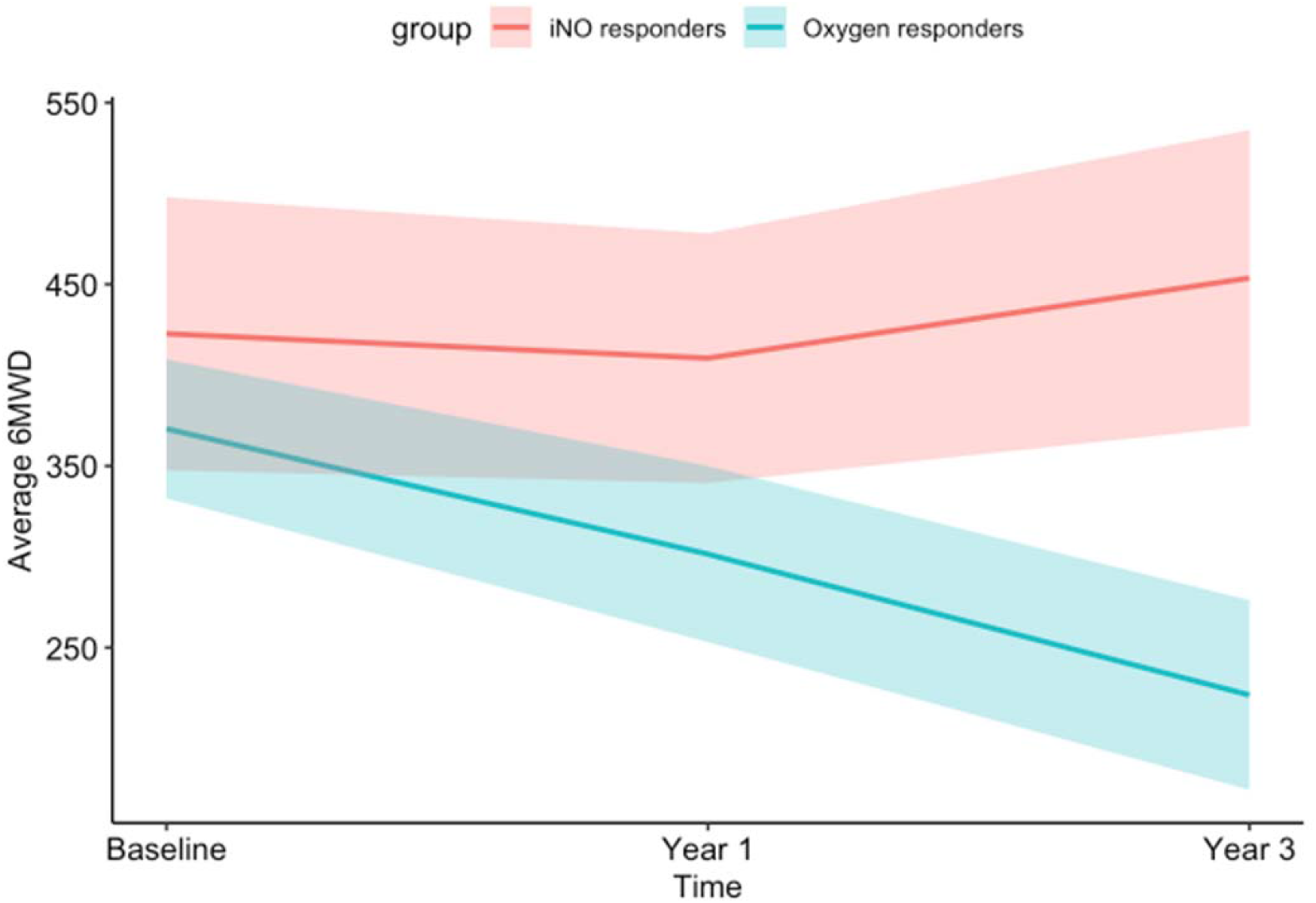
Average six-minute walk distance (6MWD) in meters at time of initiation of calcium channel blocker (Baseline) as well as 1-year and 3-year follow up for iNO Responders and Oxygen Responders. Baseline, 1-year, and 3-year 6MWD for iNO Responders were 423 m, 409 m, 454 m, respectively and for Oxygen Responders were 352 m, 292 m, and 224 m, respectively. Ribbons indicate the average 6MWD +/- 1 standard error, calculated separately from data at each time point.

## Discussion

Accurate identification of long-term CCB responders is a critical step in the evaluation of patients with PAH (7, 28). Correct identification with acute vasodilator challenge selects for patients with improved functional status and decreased mortality (4-6). Incorrect identification can lead to deleterious effects from CCB therapy including decreased cardiac output, decreased systemic mean arterial pressure, decompensated functional status, and increased mortality (6). Due to the potential hemodynamic instability caused by acute vasoresponder testing with short-acting CCBs (29-31), other agents are preferentially used. iNO is a safe, well-tolerated, and frequently utilized vasodilator agent in acute vasodilator challenge (10). iNO is delivered at variable concentrations and is often co-administered with oxygen, although there is significant variability in how oxygen is utilized (11-15). At present, there is no standardized protocol for the utilization of oxygen or the interpretation of the effects of oxygen on changes in mPAP during acute vasodilator challenge. In this study, we demonstrate that defining acute vasoresponsiveness after first adjusting for the effects of oxygenation identifies a group which has excellent long-term responsiveness to CCBs. Conversely, treating patients who primarily respond to oxygen, without a significant additional response to iNO, with calcium channel blockers may result in harm and delay appropriate treatment.

During acute vasodilator challenge, both iNO Responders and Oxygen Responders had a similar overall change in mPAP from baseline to vasodilator phase. However, when the effects of oxygen are isolated, two distinct patterns of response emerge. Amongst Oxygen Responders, over half (56.1%) of their overall decrease in mPAP occurred due to the effects of oxygenation alone. Conversely, iNO Responders had 77% of their overall reduction in mPAP from oxygen phase to vasodilator phase. The reduction in mPAP from oxygen phase to vasodilator phase was nearly 2.5 times greater in iNO Responders than in Oxygen Responders (mean decrease in mPAP of 14.4 versus 6 mmHg, respectively). The relationship between alveolar oxygen tension and pulmonary arterial vascular tone is well described (16-18) and hyperoxia has been shown to modulate mPAP (19, 20) and PVR (21) in PAH patients. Our overall cohort of 268 PAH patients had a reduction in mPAP of 3.8 mmHg, similar reduction in mPAP to other trials (19, 20). Without adjusting for the known vasodilatory effects oxygen, it is possible to incorrectly attribute vasoresponsiveness to iNO.

There was a particularly striking difference in mortality between the two cohorts. The iNO Responder cohort had no deaths in a 10-year period. Amongst Oxygen Responders, the mortality rate at 5 years exceeded 50%. Six of the 8 deaths that occurred in this cohort were from progressive right heart failure and all patients were on a multi-drug regimen for management of their PAH prior to their death. Subgroup analyses restricted to patients recommended to undergo acute vasoreactivity testing by the WSPH and ECS/ERS guidelines (26, 27) – iPAH, FPAH, and toxin-mediated PAH – yielded similar results. In this WSPH and ECS/ERS cohort, no deaths were observed in the iNO Responder group and the mortality amongst the Oxygen Responders was 100% at 5 years. This robust survival benefit observed amongst iNO Responders is in keeping with the excellent long-term prognosis described in long-term CCB responders (4, 6, 8). The accelerated mortality seen amongst Oxygen Responders is also consistent with prior cohorts of non-CCB responders who received initially CCB therapy (6). The 10-year restricted mean duration of CCB monotherapy was 6.0 in iNO Responders versus 2.8 in Oxygen Responders and nearly half of Oxygen Responders had failed CCB monotherapy after one year. Subgroup analysis of iPAH, FPAH, and toxin-mediated PAH demonstrated similar monotherapy tolerance for iNO Responders and Oxygen Responders (7.4 versus 1.5 years, respectively, *P* = 0.085), Additionally, 7 of the 23 (30.4%) Oxygen Responders had acute decompensated right heart failure within a year of initiation of CCB therapy while no iNO Responders had decompensated right heart failure within a year of CCB initiation.

Overall, iNO Responders and Oxygen Responders had similar baseline functional characteristics. However, at 1- and 3-year follow up, iNO Responders had an overall increase in their 6MWD while Oxygen Responders had a progressive decline in over this time period. At baseline, there were no significant differences in NYHA functional status. During follow up at 1 and 3 years, the proportion of iNO Responders who were NYHA I or II versus NYHA III or IV remained higher than in Oxygen Responders, though this never reached the threshold of statistical significance.

It is interesting to note that a significant difference exists between the proportion of patients with idiopathic and associated PAH amongst the two cohorts. However, there were no significant differences in survival or tolerance of monotherapy amongst patients with iPAH versus associated PAH in the Oxygen Responder cohort. Additionally, if both iNO Responders and Oxygen Responders are considered, there is a higher-than-expected rate of vasoresponsiveness amongst both associated PAH patients (9.2% vasoresponsive) and iPAH patients (14.3% vasoresponsive). If we consider iNO Responders alone, the rates are more aligned with expected values, with 1.0% of all associated PAH patients and 7.7% of all iPAH patients meeting the definition of acute vasoresponders. This demonstrates that, without correcting for oxygen, we likely decrease the specificity of acute vasodilator challenge and over diagnose acute vasoresponsiveness.

It is also notable that the study is from a pulmonary hypertension center which is located at an altitude of 5,280 feet above sea level. The phenomenon of reduced mPAP in PAH patients following administration of supplemental oxygen has been described at other altitudes. Both the average reduction in mPAP (∼ 4 mmHg) and distribution (upper quartile of responders with ≥ 6 mmHg of mPAP reduction) in our overall cohort of 268 PAH patients is consistent with other studies of PAH patients exposed to 100% FiO2 at lower altitudes (19, 20). This suggests that the response to oxygen in our cohort in neither unique nor disproportionate in magnitude or distribution to that of PAH patients at other altitudes. Additionally, our protocol of collecting baseline hemodynamic measurements at patients’ home oxygen requirement with SpO_2_ between 90%-94% should ameliorate the effects of altitude-related hypoxemia. It is also interesting to note that the rates of chronic oxygen requirement were higher amongst the iNO Responders than the Oxygen Responders, suggestive that preexisting chronic oxygen requirement is not a driving factor in oxygen responsiveness. During the course of this study, a new definition was proposed for pulmonary hypertension, using mPAP ≥ 20 mmHg as the threshold for diagnosis rather than the traditional definition of mPAP ≥ 25 mmHg (8). It remains unclear how inclusion of patients with mPAP from 20 – 24 mmHg will affect the incidence or prognosis of long-term CCB response. However, a standardized approach to the workup of PAH patients will remain imperative.

### Interpretation

The results of our study suggest that consideration of effects of oxygen during acute vasodilator challenge is critical for proper identification of long-term CCB responders. Stepwise evaluation of the effects of supplemental oxygen and iNO revealed a subset of patients with an exaggerated response to supplemental oxygenation. Failure to correct for the effects of oxygen on mPAP in the interpretation of acute vasodilator challenge with iNO selects for a patient population with inferior long-term response to CCBs, higher rates of decompensated right heart failure, and decreased survival. Those patients who demonstrate vasoresponsiveness to iNO independent from the effects oxygen have excellent long-term response to CCB monotherapy, superior functional status, and increased survival. These findings should be validated across a variety of settings. Ultimately, a standardized approach for the use of supplemental oxygen during acute vasodilator challenge with iNO is necessary.

## Data Availability

All data contained within an Excel sheet on an encrypted hard drive

## Guarantor statement

Matthew Rockstrom, MD is the guarantor of all content of the manuscript including data and analysis.

## Author contributions

All listed authors met ICMJE qualifications for authorship.

